# Accounting for early death recording in acute myocardial infarction mortality indicators based on administrative data – Observational study comparing Germany and the United States

**DOI:** 10.1101/2023.01.13.23284522

**Authors:** Ulrike Nimptsch, Thomas Mansky, Reinhard Busse

## Abstract

**Objective:** To investigate whether differences in early death recording in administrative hospital data affect the comparison of acute myocardial infarction (AMI) in-hospital mortality between Germany and the United States (U.S.), and to explore approaches to account for this issue.

**Design:** Observational cross-sectional study based on administrative hospital data. The German Diagnosis-Related Group Statistics, the U.S. National Inpatient Sample and the U.S. Nationwide Emergency Department Sample were analysed.

**Setting:** Acute inpatient and emergency department (ED) care in German and U.S. acute care hospitals from 2014 to 2019.

**Participants:** Cases with treatment for AMI were identified according to the OECD indicator “AMI 30 day in-hospital (same hospital) mortality using unlinked data (admission based)”. 1.30 million acute care hospitalizations for AMI were identified in German inpatient data and 3.88 million were estimated from U.S. inpatient data. From U.S. ED data additional 25,500 patients with a first listed diagnosis of AMI, who died before inpatient admission, were estimated.

**Primary outcome measures:** National in-hospital mortality for AMI.

**Results:** While short-duration treatments due to early death are generally recorded in German inpatient data, in U.S. inpatient data those cases are partially missing. German age- and- sex standardized in-hospital mortality was substantially higher compared to the U.S. (in 2019 7.3% vs. 4.6%). The ratio of German vs. U.S. mortality was 1.6. After consideration of ED deaths in U.S. data this ratio declined to 1.4. Exclusion of same-day stay cases in German and U.S. data led to a similar ratio.

**Conclusions:** Excluding cases with short-duration treatment from the calculation of mortality indicators might be a feasible approach to account for differences in the recording of early deaths, that might be existent in other countries as well.

**Strengths and limitations:**

- The strength of this study is the use of large national administrative data sources.
- Administrative hospital data is collected for billing purposes and differences in coding, reimbursement rules, and data collection practice may impair international comparisons.
- The deliberate assignment of ED deaths in U.S. data to a length of stay of less than one day was done for practical reasons but might not be correct in all cases.
- Possible differences in coding due to different modifications of the ICD-10 should be considered, as well as the transition from ICD-9 to ICD-10 coding that took place in the U.S. in 2015.
- The present study focused only admission-based AMI mortality, i.e., only deaths occurring in the same hospital were captured.

## Introduction

Acute myocardial infarction (AMI) is a common emergency condition in western industrialized countries. Patient survival depends on timely revascularization, preferably by means of percutaneous coronary intervention (PCI).^1^ This is why in-hospital mortality for AMI is widely used in international comparisons as an indicator of health system performance that allows conclusions to be drawn about the effectiveness of care processes.^2^

Measurement of AMI in-hospital mortality often relies on administrative hospital data, which are generated through the processes of hospital care. For instance, the AMI mortality indicators of the OECD Health Care Quality Indicators (HCQI) programme are based on administrative hospital data from the respective member states.^3^

The HCQI comparison of admission-based age-and-sex standardized thirty-day mortality for AMI shows considerable variation across countries: While in the year 2017 the United States (U.S.) rate of 5.0% was below the OECD average of 6.9%, in Germany 8.5% of AMI patients died after admission.^4^ These figures might reflect subpar AMI care in Germany and there might be potential for improvement.^5^ However, compared to other countries, the German health system is characterized by high hospital capacity and easy access to immediate hospital services.^4, 6^ In 2019 more than 80% of AMI patients in Germany received coronary angiography or PCI in the first treating hospital.^7^

As mentioned by the OECD, differences in administrative data-based AMI mortality figures between countries might not solely reflect differences in health system performance, but as well differences in length of stay, transfers to other hospitals, or disease severity.^4^ Yet, there might be another source of bias which has not been comprehensively explored, namely, whether short treatments due to early death after arrival to the hospital are completely captured in inpatient databases. This may vary in different countries, depending on billing practices within the respective health system. Regarding AMI mortality this issue is of relevance, since fatal events may occur in the emergency department (ED) before the patient is admitted to a specialized hospital ward. A German clinical registry reported that almost one third of deaths in hospitalized patients diagnosed with AMI occurred during the first 24 hours after onset of symptoms.^8^

In Germany, treatments of patients who died shortly after arrival to the hospital are normally billed as inpatient cases via the all-payer German Diagnosis Related Groups System. Although this practice has been subject to legal disputes between hospitals and statutory health insurance funds in the past, it has been confirmed by German case-law several times.^9, 10^ This implies that short-duration treatments due to early death are virtually completely recorded in German inpatient data. A recent study of combined ED, inpatient, and outpatient data of 16 large German hospitals found that the percentage of early fatalities with outpatient billing was less than 1%.^11^

In the U.S., treatments of patients who died in the ED might be billed as outpatient if they were not admitted to a hospital ward before death. Regarding Medicare beneficiaries the Centers for Medicare & Medicaid Services (CMS) specified in 2013 that cases should be billed as inpatient if the patient is expected to require a hospital stay that crossed two midnights. This also includes stays in which this expectation is supported, but the length of the actual stay was less than two midnights due to death.^12^ However, this rule seems to be subject to debate, in particular regarding the case of AMI.^13^ Moreover, a majority of the U.S. population receives their coverage from private health insurance,^14^ and billing practices regarding inpatient or outpatient payment may vary by provider.^15^ Therefore, in contrast to Germany, treatments for AMI followed by early death might be less completely recorded in U.S. inpatient data.

The aim of this study was to investigate whether early death recording affects the comparison of AMI in-hospital mortality between Germany and the U.S., and to explore approaches to account for this issue by modifying mortality indicator definitions. Beyond that, the analysis provides a cross-country comparison of AMI morbidity and patterns of care.

## Methods

### Study design

Within an observational cross-sectional study design national in-hospital mortality for AMI was compared between Germany and the U.S. Different modifications to the composition of mortality figures were applied to administrative data from acute inpatient and emergency department care.

### Data

For Germany, the Diagnosis-Related Group Statistics (DRG Statistics) from 2014 to 2019 were analysed. The DRG Statistics are a complete all-payer database comprising data records of all inpatient stays in all German acute care hospitals, except for psychiatric and psychosomatic treatment. The data contain principal and secondary diagnoses that are coded according to the German modification of the International Classification of Diseases (ICD-10-GM). Procedures are coded according to the German procedure coding system (Operationen-und Prozedurenschlüssel, OPS). Information on sex, age, source of admission, discharge disposition, and length of stay are also included. The data were provided by the Research Data Centre of the German Federal Statistical Office and were accessed via remote execution.^16^

For the U.S., the National Inpatient Sample (NIS) and the Nationwide Emergency Department Sample (NEDS) of the Health-Care Cost and Utilization Project (HCUP) from 2014 to 2019 were analysed. NIS is an all-payer sample of inpatient stays from all hospitals participating in HCUP, covering about 7 million inpatient stays per year.^17^ NEDS is an all-payer ED database, covering about 30 million ED visits with or without inpatient admission per year.^18^ Diagnosis coding in both datasets changed from ICD-9 Clinical Modification (ICD-9-CM) to ICD-10-CM in the year 2015. The first listed diagnosis in an inpatient record or an ED record represents the principal diagnosis. Procedures are coded according to the ICD-9 procedure coding system (PCS), or ICD-10-PCS, respectively. In NEDS data, procedures undertaken during ED visits without inpatient admission are coded according to the Clinical Classifications Software (CCS) services and procedures classification. Both datasets contain information on sex, age, and discharge disposition. Information on length of stay and source of admission are only available for inpatient cases. Both, NIS and NEDS represent approximately 20% of inpatient stays or, respectively, ED visits in U.S. hospitals. Weights are available to calculate estimates for the entire U.S. population. NIS and NEDS data were provided by the HCUP Central Distributor.

### Cases

In the German DRG Statistics, as well as in the U.S. NIS, inpatient cases with treatment for AMI were identified by applying the inclusion criteria of the OECD indicator AC2 “AMI 30 day in-hospital (same hospital) mortality using unlinked data (admission based)”. The indicator covers hospital admissions for acute care with a principal diagnosis of AMI of patients aged 15 years and older. All admissions (including day cases) are to be counted in the denominator including cases transferred-in from another hospital, as well as cases transferred-out to another hospital. The numerator of this indicator comprises deaths in the same hospital that occurred within 30 days of the admission date.^19^ Additionally, the OECD indicator definition requires a restriction to cases with emergency treatment. However, this requirement could not be met, as German data allows no valid separation of emergency from elective, or non-urgent status. Although German data contain a flag for ‘emergency’ admission, this flag actually identifies cases without referral by a resident physician. As access to hospitals in Germany is not restricted, this flag rather depicts the administrative mode of access than clinical emergency. On the other hand, U.S. NIS data contain a flag for elective admission. Initial analysis of NIS data revealed that the proportion of AMI cases coded as elective was less than 5% and in-hospital mortality of those cases (4.3%) was only slightly deviant from the mortality of the whole AMI inpatient population (4.6%). Therefore, it was decided to keep all cases in the analysis of German and U.S. data, regardless of emergency or elective status.

Aiming to identify AMI cases that are not recorded in U.S. inpatient data ED deaths without inpatient admission of patients with a first listed diagnosis of AMI aged 15 years and older were extracted from NEDS data. AMI cases transferred-out from the ED to another hospital were not considered, as those cases will likely appear in inpatient data after admission to the designated hospital. Cases that were released alive from the ED without inpatient admission were not considered, because in those cases the diagnosis of AMI might rather represent a ruled-out diagnosis.^20^ Details of inclusion and exclusion criteria are given in supplementary material 1. Case selection flow is displayed in supplementary material 2.

### Analysis

While German data represent a full sample of all inpatient cases in German hospitals, national numbers of U.S. inpatient cases and ED visits were estimated by using the respective weights provided by HCUP.

Characteristics of cases were analysed descriptively. Definitions of presented variables are displayed in supplementary material 1. German and U.S. population-based AMI case rates were calculated and standardized by 5-year age groups and sex according to the OECD 2010 standard population.^21^

To assess differences in patterns of care and mortality, percentage distributions of cases and deaths were stratified by length of stay. As no time stamp is available in U.S. data and the validity of time stamps in German data is uncertain,^22^ length of stay was determined by admission date. Cases with a length of stay of 0 days (i.e., discharge date - admission date = 0) were determined as same-day stays. As in U.S. data no information on length of stay is available for ED visits without inpatient admission, ED death cases were assigned to a length of stay less than one day and thus determined as same-day stays.

Age-and-sex specific mortality rates were displayed for all AMI cases as well as for the subgroup of cases with transmural/ST-elevation myocardial infarction (STEMI).

AMI in-hospital mortality was calculated for each year of observation. German mortality figures were standardized by 5-year age groups and sex according to the U.S. AMI disease population of the respective calendar year. According to the definition of the OECD indicator mortality figures refer to cases aged 45 years and above.

To assess the possible bias resulting from differences in the recording of early deaths, deliberate modifications were applied to the composition of mortality figures. In a first step U.S. mortality figures were recalculated after adding ED deaths without admission to the denominator and the nominator. In a second step German and U.S. mortality figures were recalculated after excluding same-day stay cases from the denominator and the nominator. The analyses were repeated for the subgroup of STEMI cases. Differences between German and U.S. mortality figures were expressed as ratios.

The analyses were conducted using SAS Version 9.4 (SAS Institute Inc., Cary, NC, U.S.A). Reporting adheres to the RECORD (REporting of studies Conducted using Observational Routinely-collected health Data) Statement.^23^

### Patient and Public Involvement

There was no patient or public involvement.

## Results

In Germany, 1.30 million hospitalizations for acute myocardial infarction were observed from 2014 to 2019. In the U.S., 3.88 million AMI hospitalizations were estimated during this time span. After consideration of about 25,500 cases with a first listed diagnosis of AMI who died in the ED without inpatient admission, the estimated number of AMI cases in the U.S. was 3.90 million. After age-sex standardization average annual rates per 100,000 population in Germany (235) and the U.S. (236) were similar (table 1).

**Table 1.** Characteristics of cases treated for acute myocardial infarction, accumulated data of 2014 to 2019.

|  | Germany<br>(full sample) | United States<br>(estimates weighted to national average) |  |  |
| --- | --- | --- | --- | --- |
|  | DRG statistics | National<br>Inpatient Sample<br>(NIS) | Nationwide<br>Emergency<br>Department Sample<br>(NEDS) | NIS and NEDS<br>combined |
|  | *Inpatient treatment*<br>*(including early*<br>*deaths)* | *Inpatient treatment* | *ED death without*<br>*admission* | *Inpatient treatment*<br>*or ED death without*<br>*admission* |
| Cases with a principal or first listed diagnosis of AMI N (%) | 1,300,718 (100.0) | 3,875,610 (100.0) | 25,598 (100.0) | 3,901,208 (100.0) |
| Per 100,000 population per year | 262.8 | 199.3 | 1.3 | 200.6 |
| Per 100,000 population per year, age-and-sex standardized <sup>a</sup> | 235.1 | 234.5 | 1.6 | 236.1 |
| Female N (%) | 433,530 (33.3) | 1,467,585 (37.9) | 10,553 (41.2) | 1,478,138 (37.9) |
| Age >= 65 years N (%) | 847,114 (65.1) | 2,209,870 (57.0) | 17,860 (69.8) | 2,227,729 (57.1) |
| Age Median (P25 - P75) | 72 (60 - 80) | 67 (57 - 77) | 72 (62 - 83) | 67 (57 - 77) |
| Transmural/ST-elevation AMI N (%) | 422,633 (32.5) | 1,001,485 (25.8) | 14,100 (55.1) | 1,015,585 (26.0) |
| Cardiogenic shock N (%) | 85,258 (6.6) | 240,130 (6.2) | 2,427 (9.5) | 242,557 (6.2) |
| Resuscitation (%) | 71,798 (5.5) | 71,640 (1.8) | 13,313 (52.0) | 84,953 (2.2) |
| Percutaneous coronary intervention N (%) | 766,497 (58.9) | 1,835,440 (47.4) | 1,714 (6.7) | 1,837,154 (47.1) |
| Coronary artery bypass graft N (%) | 71,597 (5.5) | 338,870 (8.7) | <=10 (0.0) | 338,875 (8.7) |
| Transferred-in from other acute care hospital N (%) | 175,698 (13.5) | 695,220 (17.9) | n/a | 695,220 (17.8) |
| Treated in emergency department N (%) | n/a | 2,811,935 (72.6) | 25,598 (100.0) | 2,837,533 (72.7) |
| Same-day stay N (%) <sup>b</sup> | 82,033 (6.3) | 118,010 (3.0) | n/a | 143,608 (3.7) |
| Length of stay Median (P25 - P75) | 6 (3 - 9) | 2 (1 - 5) | n/a | 2 (1 - 5) |
| Transferred-out to other acute care hospital N (%) | 215,728 (16.6) | 298,680 (7.7) | n/a | 298,680 (7.7) |
| In-hospital death N (%) | 107,741 (8.3) | 182,340 (4.7) | 25,598 (100.0) | 207,938 (5.3) |
| In-hospital death within 30 days N (%) | 104,831 (8.1) | 179,730 (4.6) | 25,598 (100.0) | 205,328 (5.3) |

AMI: acute myocardial infarction; ED: emergency department; n/a: not available. <sup>a</sup> Directly standardized by sex and 5-year age-groups according to the 2010 OECD standard population, age >=15. <sup>b</sup> As in U.S. NEDS data no information on length of stay is available for ED visits without inpatient admission, ED deaths without admission were assigned to a length of stay <1 day (same-day stay).

Over time, age-and-sex standardized rates declined in Germany from 246 to 225, while the U.S. rate of 234 in 2014 was transiently elevated in 2016 and 2017, followed by a subsequent decline to 232 cases per 100,000 population in 2019 (see supplementary material 3 for characteristics by year).

Compared to the U.S., the percentage of females was lower in Germany (33% vs. 38%) and median age was higher (72 vs. 67). In Germany, the percentage of STEMI was higher (33% vs. 26%), and a higher percentage of cases received PCI (59% vs. 47%) while coronary artery bypass surgery was less frequently performed (6% vs. 9%). Cardiopulmonary resuscitation was more often coded in Germany (5.5%) than in the U.S. (2.2% after consideration of ED deaths, table 1).

The percentage distribution of cases by length of stay (LOS) revealed different patterns. While in the U.S. there was a marked peak at a LOS of two days, in Germany LOS was more broadly distributed around five days (figure 1). The percentage distribution of deaths showed that in Germany more than one quarter of deaths of patients diagnosed with AMI occurred at the admission date. In U.S. data this proportion was quite similar after considering ED deaths without admission as same-day stays (figure 1).

**Figure 1.**
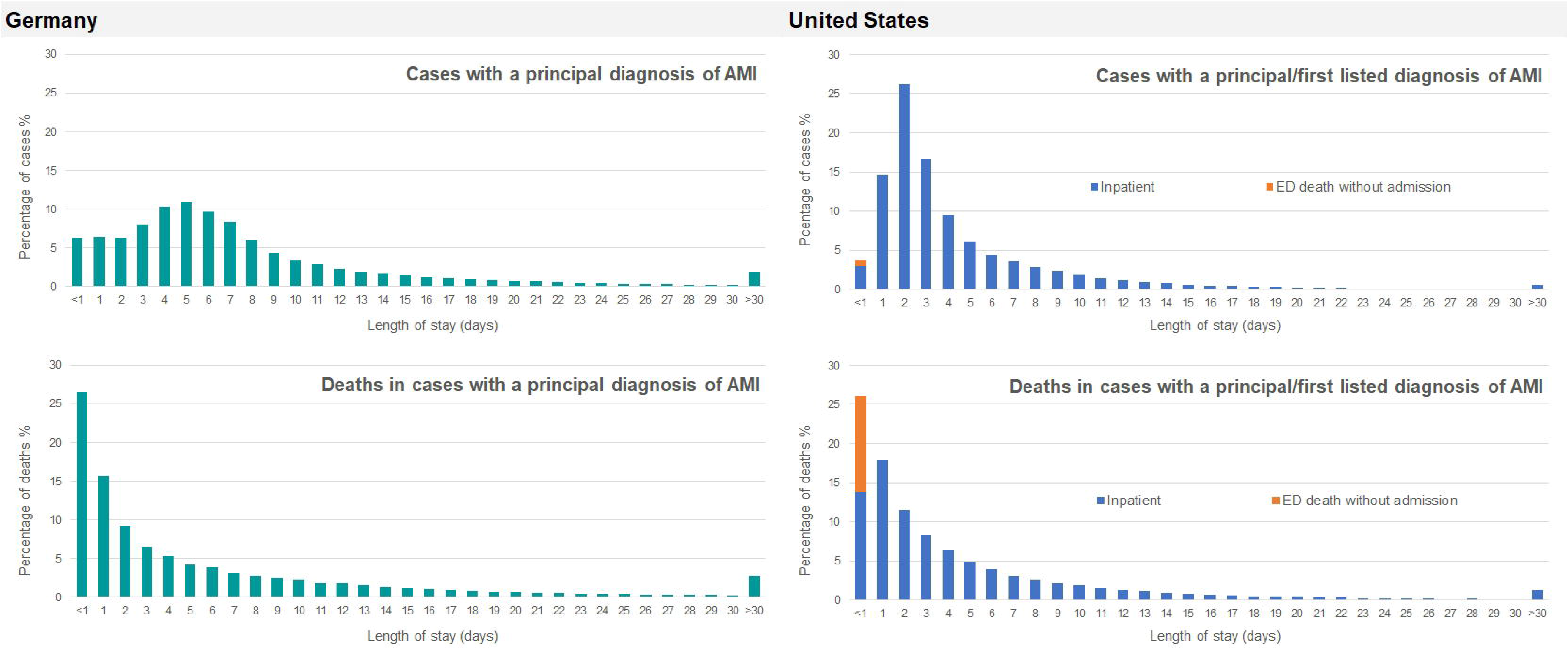
Percentage distribution of cases treated for acute myocardial infarction and percentage distribution of deaths by length of stay, accumulated data of 2014 to 2019. AMI: acute myocardial infarction; ED: emergency department. Note: As in US NEDS data no information on length of stay is available for ED visits without inpatient admission, ED deaths without admission were assigned to a length of stay <1 day (same-day stay).

Age-and-sex specific mortality rates revealed that mortality in Germany was substantially higher throughout all age groups. Regarding the whole AMI population those differences were most pronounced in women aged 85 years and above. Considering ED deaths in U.S. data reduced, but not diminished age-and-sex specific mortality differences. After restriction to the subgroup of cases with STEMI the mortality differences between Germany and the U.S. were smaller, but still existent in most age-sex groups (figure 2).

**Figure 2.**
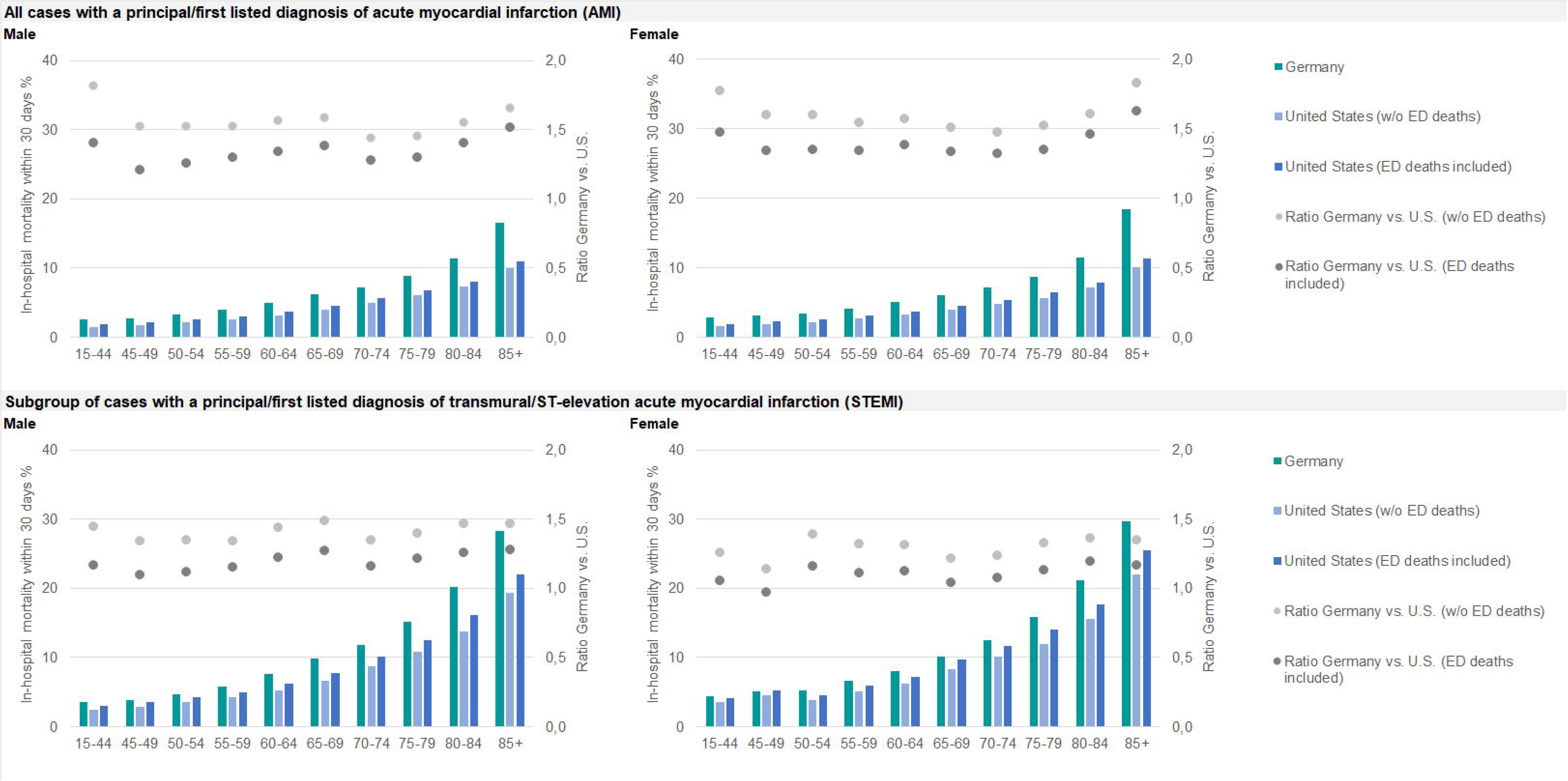
30-day mortality in the same hospital of cases treated for acute myocardial infarction stratified by sex and age groups, accumulated data of 2014 to 2019. ED: emergency department.

The comparison of mortality figures according to the OECD indicator definition showed higher German age-and-sex standardized in-hospital mortality compared to the U.S. (in 2019 7.3% vs. 4.6%, figure 3). In both countries a decline was visible from 2014 to 2019. During this time span the ratio of German vs. U.S. mortality was stable at 1.6. After consideration of ED deaths in U.S. data this ratio declined to 1.4. Exclusion of cases with same-day stay led to a similar ratio (table 2, figure 3).

**Figure 3.**
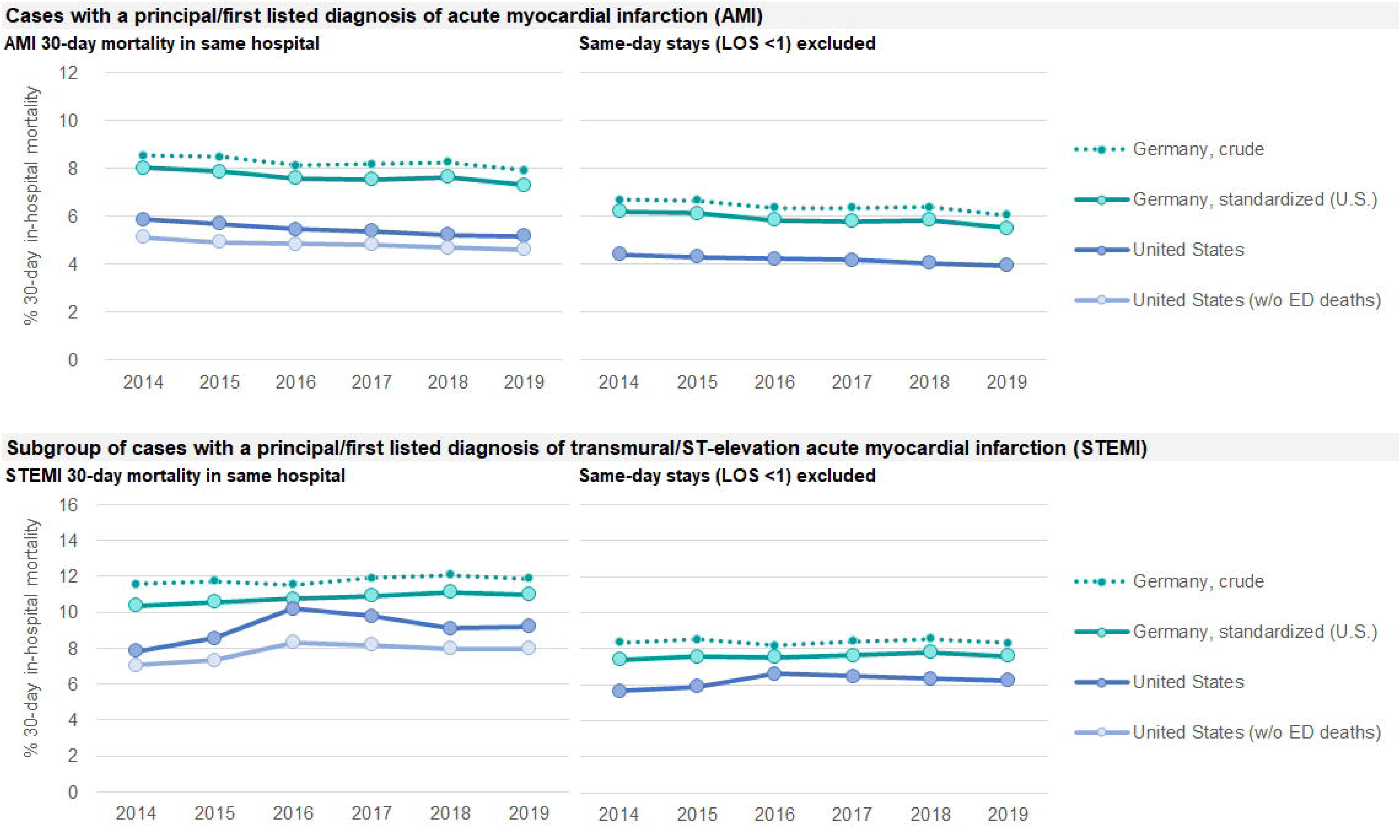
30-day mortality in the same hospital of cases treated for acute myocardial infarction, 2014 to 2019. LOS: Length of stay. Note: According to the definition of the OECD indicator AC2 “AMI 30 day in-hospital (same hospital) mortality using unlinked data (admission based)” all figures refer to cases aged 45 years and above. German figures were standardized to the US disease population of the respective calendar year.

**Table 2.** 30-day mortality in the same hospital of cases treated for acute myocardial infarction, 2014 to 2019.

|  | 2014 | 2015 | 2016 | 2017 | 2018 | 2019 |
| --- | --- | --- | --- | --- | --- | --- |
| **All cases with acute myocardial infarction (AMI)** |  |  |  |  |  |  |
| **AMI 30-day mortality in same hospital** |  |  |  |  |  |  |
| Germany (standardized to United States disease population) % | 8.0 | 7.8 | 7.6 | 7.5 | 7.6 | 7.3 |
| United States (inpatient data only) % | 5.1 | 4.9 | 4.8 | 4.8 | 4.7 | 4.6 |
| *Ratio Germany vs. United States* | 1.6 | 1.6 | 1.6 | 1.6 | 1.6 | 1.6 |
| **AMI 30-day mortality in same hospital, ED deaths considered in US data** |  |  |  |  |  |  |
| Germany (standardized to United States disease population) % | 8.0 | 7.8 | 7.6 | 7.5 | 7.6 | 7.3 |
| United States (including ED deaths without admission) % | 5.9 | 5.7 | 5.4 | 5.4 | 5.2 | 5.2 |
| *Ratio Germany vs. United States* | 1.4 | 1.4 | 1.4 | 1.4 | 1.5 | 1.4 |
| **AMI 30-day mortality in same hospital, same-day stays (LOS <1) excluded** |  |  |  |  |  |  |
| Germany (standardized to United States disease population) % | 6.2 | 6.1 | 5.8 | 5.8 | 5.8 | 5.5 |
| United States (inpatient data only) % | 4.4 | 4.3 | 4.2 | 4.2 | 4.0 | 3.9 |
| *Ratio Germany vs. United States* | 1.4 | 1.4 | 1.4 | 1.4 | 1.4 | 1.4 |
| **Subgroup of cases with transmural/ST-elevation (STEMI) myocardial infarction** |  |  |  |  |  |  |
| **Transmural/STEMI 30-day mortality in same hospital** |  |  |  |  |  |  |
| Germany (standardized to United States disease population) % | 10.3 | 10.5 | 10.7 | 10.8 | 11.1 | 10.9 |
| United States (inpatient data only) % | 7.1 | 7.3 | 8.3 | 8.2 | 8.0 | 8.0 |
| *Ratio Germany vs. United States* | 1.5 | 1.4 | 1.3 | 1.3 | 1.4 | 1.4 |
| **Transmural/STEMI 30-day mortality in same hospital, ED deaths considered in US data** |  |  |  |  |  |  |
| Germany (standardized to United States disease population) % | 10.4 | 10.6 | 10.8 | 10.9 | 11.1 | 11.0 |
| United States (including ED deaths without admission) % | 7.8 | 8.6 | 10.2 | 9.8 | 9.1 | 9.2 |
| *Ratio Germany vs. United States* | 1.3 | 1.2 | 1.1 | 1.1 | 1.2 | 1.2 |
| **Transmural/STEMI 30-day mortality in same hospital, same-day stays (LOS <1) excluded** |  |  |  |  |  |  |
| Germany (standardized to United States disease population) % | 7.4 | 7.6 | 7.5 | 7.6 | 7.8 | 7.6 |
| United States (inpatient data only) % | 5.6 | 5.9 | 6.6 | 6.5 | 6.3 | 6.2 |
| *Ratio Germany vs. United States* | 1.3 | 1.3 | 1.1 | 1.2 | 1.2 | 1.2 |

ED: emergency department; LOS: length of stay. Note: According to the definition of the OECD indicator AC2 “AMI 30 day in-hospital (same hospital) mortality using unlinked data (admission based)” all figures refer to cases aged 45 years and above. German figures were standardized to the U.S. disease population of the respective calendar year.

In the subgroup of STEMI cases German age-and-sex standardized in-hospital mortality was higher than in the U.S. (in 2019 10.9% vs. 8.0%, figure 3). A slight increase of in-hospital mortality was observed over time in both countries. In U.S. data a transient elevation of mortality was found in 2016, the year after transition from ICD-9-CM to ICD-10-CM (figure 3). In 2019 the ratio of German vs. U.S. mortality was 1.4. After consideration of ED deaths in U.S. data this ratio declined to 1.2. Exclusion of cases with same-day stay led to a similar ratio (table 2, figure 3).

## Discussion

In Germany as well as in the U.S., more than one quarter of deaths of patients diagnosed with AMI occurred within 24 hours after arrival to the hospital, or in the ED before inpatient admission. While short-duration treatments due to early death are generally recorded in German inpatient data, in U.S. inpatient data those cases are partially missing.

Consequently, the consideration of ED deaths without inpatient admission in U.S. data resulted in higher mortality figures in contrast to using only inpatient data. However, although the difference between German and U.S. mortality declined after accounting for ED deaths in U.S. data German AMI mortality remained substantially higher.

### Strengths and limitations of this study’

The strength of this study is the analysis of complete national administrative hospital data from Germany and large representative national samples of administrative hospital and ED data from the U.S. However, several limitations should be considered. First, administrative hospital data is collected for billing purposes and differences in coding, reimbursement rules, and data collection practice may impair international comparisons.^24^ One of those issues is the recording of early deaths which was subject of this study. Second, the deliberate assignment of ED deaths in U.S. data to a length of stay of less than one day was done for practical reasons. However, it is possible that this assumption might not be correct in all cases. In this context, it should also be considered that length of stay was determined based on the admission date instead of exact admission time. This might have caused an incomplete separation of cases with an overnight stay of stay less than 24 hours. Third, possible differences in coding due to different modifications of the ICD-10 should be considered. While the German Modification bases its classification of AMI on anatomic criteria (transmural vs. non-transmural infarction), the Clinical Modification used in the U.S. allows the electrophysiological distinction between ST-elevation and non-ST-elevation infarctions. As well, the transition from ICD-9 to ICD-10 coding that took place in the U.S. in 2015 should be considered. Finally, one should note that the present study focused only admission-based AMI mortality, i.e., only deaths occurring in the same hospital were considered. The OECD recommends calculating rather patient-based mortality indicators which consider all deaths that occurred after initial hospital admission, whether in the same or another hospital or outside hospital. While this method is considered more robust, it requires a unique patient identifier to link the data across the relevant datasets.^19^ However, the studied data do not contain a patient identifier.

The results of this study suggest that international comparisons of AMI mortality figures can be biased by international differences in the recording of early deaths, as different reimbursement systems may cause a different administrative handling of short stay cases. Consequently, such cases may not completely appear in inpatient data. Therefore, the approach of excluding cases with a short duration of treatment from the calculation of mortality figures was explored. The rationale was, focusing only those cases that are most likely completely recorded in inpatient data of each country might allow more reliable comparisons. In fact, the ratio between German and U.S. AMI mortality was quite similar after exclusion of same-day cases, compared to the ratio after including ED deaths in U.S. data.

This study investigated the case of Germany and the U.S. because sufficient data is available from both countries. Yet, differences in the recording of early deaths might be existent in other countries as well, in one way or another. The HCQI expert group stated that indicators for international comparison should be defined in a way that indicator results reflect issues in quality of care rather than differences in non-quality of care reasons, such as data collection methodologies. On the other hand, indicator definitions should also be internationally feasible, i.e., data should be derived without substantial additional resources.^25^ The approach of excluding same-day cases from the calculation of mortality figures might be a feasible way to account for international differences in the recording of early deaths without additional use of ED data. This approach might also reduce possible bias resulting from differences in the frequency of early transfers to another hospital, or differences in diagnostic accuracy which might be more existent in cases with a short duration of treatment. However, additional research on data from other countries is needed to confirm the appropriateness of this approach.

In addition to international comparisons, it should also be mentioned that incomplete coverage of early deaths in inpatient data may affect the national use of AMI mortality in inpatient quality indicator systems. This, in turn, may be relevant for value-based reimbursement methods.

Another finding of the present study is that German AMI mortality was substantially higher as compared to the U.S., even after accounting for early death recording. In line with these results, high AMI in-hospital mortality in Germany has been reported by international studies in the past,^26^ as well as rather low AMI mortality in the U.S.^27^ The present study revealed that AMI cases in Germany were older, less often female and were more often diagnosed with STEMI compared to U.S. AMI cases. Demographic differences in the disease populations were corrected by age-and-sex standardization. Possibly, the additional consideration of type of AMI in the calculation of mortality figures might further enhance comparability, since the mortality difference between Germany and the U.S. in the subgroup of STEMI cases was smaller than in the whole AMI population.

The descriptive analysis also revealed differences in patterns of care, such as a higher percentage of treatment with PCI and lower percentage of treatment with CABG in Germany compared to the U.S. Some of the findings might also indicate differences in AMI morbidity between both countries. In Germany, a twofold higher resuscitation rate was observed, that might – along with the higher percentage of STEMI cases – reflect a higher morbidity of the German AMI population.

## Conclusion

Indicators of health care performance allow to identify potential for improvement in individual countries and can meaningfully support health policy decisions. Because of their availability and completeness administrative hospital data might be the most suitable data source for international comparisons. However, defining indicators that reflect quality of care while being robust against non-quality-of-care related differences is challenging. Excluding cases with short-duration treatment from the calculation of AMI mortality figures might be a feasible approach to correct for differences in the recording of early deaths, that might be existent in other countries as well.

## Data Availability

German inpatient data (DRG statistics) are available for research purposes from the Research Data Centre of the Federal Statistical Office upon application. U.S. inpatient (NIS) and emergency department (NEDS) data are available from the HCUP Central Distributor upon application. Aggregated data are available from the corresponding author upon request.

## Contributorship statement

Ulrike Nimptsch, Thomas Mansky, and Reinhard Busse designed the study. Ulrike Nimptsch conducted the analysis, interpreted the data, and drafted the manuscript. Thomas Mansky and Reinhard Busse contributed to the interpretation of data and to revising the manuscript critically for important intellectual content. All authors gave final approval of the version to be published and agree to be accountable for all aspects of the work in ensuring that questions related to the accuracy or integrity of any part of the work are appropriately investigated and resolved.

## Acknowledgement

The authors thank Claire Schippers for contributing to the elaboration of the research question.

## Ethics approval

As this study relies on anonymized administrative data it was not subject to an institutional review.

