## Supplementary material for "Accounting for early death recording in acute myocardial infarction mortality indicators based on administrative data – Observational study comparing Germany and the United States"

#### Supplementary material 1 Definition of study variables

| | Germany | | United States | |
|---|---|---|---|---|
| | DRG statistics | | National Inpatient Sample (NIS) and Nationwide Emergency Department Sample (NEDS) | |
| | **Inclusion** | **Exclusion** | **Inclusion** | **Exclusion** |
| **Unit of analysis** | | | | |
| Treatment for acute myocardial infarction | PD ICD-10-GM I21 I22 | Age < 15 | PD ICD-10-CM I21 I22; PD ICD-9-CM 410 | Age < 15 |
| **Clinical presentation and severity** | | | | |
| Transmural/ST-elevation myocardial infarction (STEMI) | PD ICD-10-GM I210 I211 I212 I213 | | PD ICD-10-CM I210 I211 I212 I213;<br>PD ICD-9-CM 4100 4101 4102 4103 4104 4105 4106 4108 | |
| Cardiogenic shock | SD ICD-10-GM R570 | | SD ICD-10-CM R570; SD ICD-9-CM 78551 | |
| Resuscitation | OPS 8771 | | ICD-9-CM 9960; ICD-10-PCS 5A12012;<br>CCS 92950 | |
| **Revascularization** | | | | |
| Percutaneous coronary intervention (without coronary surgery) | OPS 88370 88371 8837288375 88376 88377 88378 88379 8837k 8837m 8837p 8837q 8837t 8837u 8837v 8837w 88399 883d0 883d1 883d2 | OPS 5361 5362 5363 | ICD-9-CM 0066 3601 3602 3605<br>ICD-10-PCS 02703 02704 02713 02714 02723 02724 02733 02734; CCS 45 | ICD-9-CM 3610 3611 3612 3613 3614 3615 3616 3617 3619; ICD-10-PCS 02100 02110 02120 02130; CCS 44 |
| Coronary artery bypass graft | OPS 5361 5362 5363 | | ICD-9-CM 3610 3611 3612 3613 3614 3615 3616 3617 3619; ICD-10-PCS 02100 02110 02120 02130; CCS 44 | |

PD: principal or first listed diagnosis, SD: secondary diagnosis, ICD: international classification of diseases, GM: German modification, CM: clinical modification, OPS: German procedure coding system, PCS: procedure coding system, CCS: Clinical Classifications Software services and procedures classification.

### Supplementary material 2 Selection of study population, accumulated data of 2014 to 2019

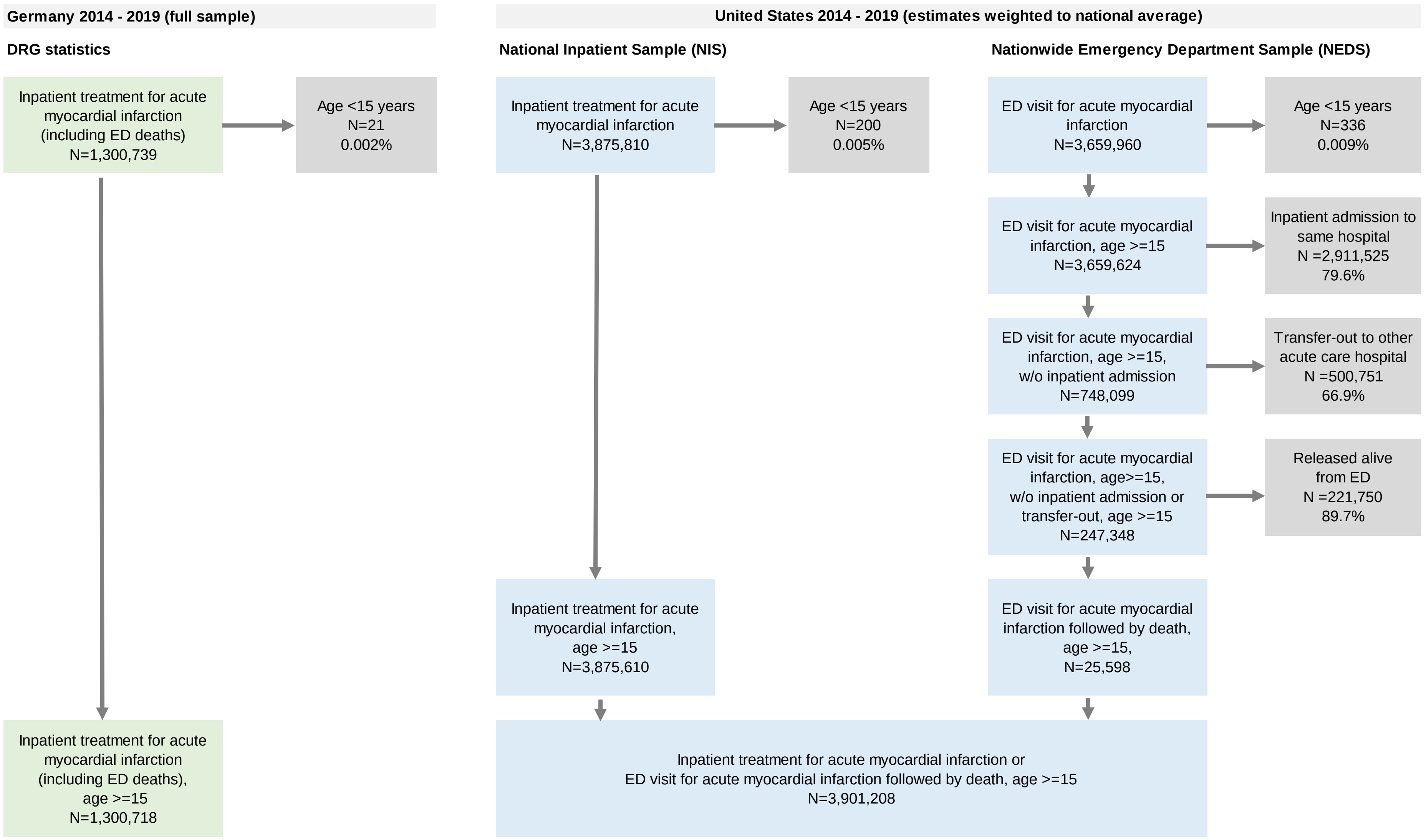

**Supplementary material 3 Characteristics of cases treated for acute myocardial infarction by year, 2014-2019**

| | Year | Germany (full sample) | United States (estimates weighted to national average) | | |
|---|---|---|---|---|---|
| | | DRG statistics | National Inpatient Sample (NIS) | Nationwide Emergency Department Sample (NEDS) | NIS and NEDS combined |
| | | *Inpatient treatment (including ED deaths)* | *Inpatient treatment* | *ED death without admission* | *Inpatient treatment or ED death without admission* |
| Cases with a principal/first listed diagnosis of AMI (age>=15) N | 2014 | 219,546 | 608,760 | 4,790 | 613,550 |
| | 2015 | 219,248 | 629,890 | 4,981 | 634,871 |
| | 2016 | 219,229 | 651,189 | 4,162 | 655,351 |
| | 2017 | 217,715 | 662,075 | 3,924 | 665,998 |
| | 2018 | 212,350 | 658,545 | 3,763 | 662,309 |
| | 2019 | 212,630 | 665,150 | 3,978 | 669,128 |
| Cases per 100,000 population (age-and-sex standardized) [a] | 2014 | 270.4 (245.6) | 190.9 (232.1) | 1.5 (1.8) | 192.4 (233.9) |
| | 2015 | 266.8 (241.7) | 196.0 (235.3) | 1.5 (1.9) | 197.5 (237.1) |
| | 2016 | 265.7 (238.9) | 201.5 (239.0) | 1.3 (1.5) | 202.8 (240.5) |
| | 2017 | 263.0 (234.6) | 203.3 (237.9) | 1.2 (1.4) | 204.5 (239.3) |
| | 2018 | 255.8 (226.8) | 201.3 (232.5) | 1.2 (1.3) | 202.4 (233.8) |
| | 2019 | 255.7 (224.5) | 202.6 (231.0) | 1.2 (1.4) | 203.9 (232.4) |
| Female N (%) | 2014 | 74,970 (34.1) | 232,325 (38.2) | 1,996 (41.7) | 234,322 (38.2) |
| | 2015 | 74,132 (33.8) | 240,905 (38.2) | 2,017 (40.5) | 242,922 (38.3) |
| | 2016 | 73,333 (33.5) | 247,450 (38.0) | 1,686 (40.5) | 249,136 (38.0) |
| | 2017 | 72,217 (33.2) | 251,830 (38.0) | 1,673 (42.6) | 253,503 (38.1) |
| | 2018 | 69,620 (32.8) | 247,125 (37.5) | 1,523 (40.5) | 248,648 (37.5) |
| | 2019 | 69,258 (32.6) | 247,950 (37.3) | 1,658 (41.7) | 249,608 (37.3) |
| Age >= 65 years N (%) | 2014 | 143,810 (65.5) | 344,140 (56.5) | 3,198 (66.8) | 347,338 (56.6) |
| | 2015 | 143,751 (65.6) | 357,625 (56.8) | 3,526 (70.8) | 361,151 (56.9) |
| | 2016 | 142,557 (65.0) | 371,115 (57.0) | 2,900 (69.7) | 374,015 (57.1) |
| | 2017 | 141,391 (64.9) | 377,885 (57.1) | 2,720 (69.3) | 380,605 (57.1) |
| | 2018 | 137,583 (64.8) | 376,615 (57.2) | 2,626 (69.8) | 379,241 (57.3) |
| | 2019 | 138,022 (64.9) | 382490 (57.5) | 2889 (72.6) | 385379 (57.6) |
| Age Median (P25 - P75) | 2014 | 72 (60 - 80) | 66 (56 - 77) | 72 (60 - 83) | 66 (56 - 77) |
| | 2015 | 72 (60 - 80) | 67 (57 - 77) | 73 (62 - 84) | 67 (57 - 77) |
| | 2016 | 72 (60 - 80) | 67 (57 - 77) | 72 (62 - 83) | 67 (57 - 77) |
| | 2017 | 71 (60 - 80) | 67 (57 - 77) | 71 (61 - 82) | 67 (57 - 77) |
| | 2018 | 71 (60 - 80) | 67 (57 - 77) | 72 (62 - 82) | 67 (57 - 77) |
| | 2019 | 71 (60 - 80) | 67 (57 - 76) | 74 (63 - 84) | 67 (57 - 76) |

### Supplementary material 3 (continued)

| | Year | Germany (full sample) | United States (estimates weighted to national average) | | |
|---|---|---|---|---|---|
| | | DRG statistics | National Inpatient Sample (NIS) | Nationwide Emergency Department Sample (NEDS) | NIS and NEDS combined |
| | | *Inpatient treatment (including ED deaths)* | *Inpatient treatment* | *ED death without admission* | *Inpatient treatment or ED death without admission* |
| Transmural/ST-elevation AMI N (%) | 2014 | 74,702 (34.0) | 151,285 (24.9) | 1,226 (25.6) | 152,511 (24.9) |
| | 2015 | 71,870 (32.8) | 158,755 (25.2) | 2,061 (41.4) | 160,816 (25.3) |
| | 2016 | 70,484 (32.2) | 176,415 (27.1) | 3,560 (85.5) | 179,975 (27.5) |
| | 2017 | 69,383 (31.9) | 173,485 (26.2) | 2,962 (75.5) | 176,447 (26.5) |
| | 2018 | 68,734 (32.4) | 168,550 (25.6) | 2,024 (53.8) | 170,574 (25.8) |
| | 2019 | 67,460 (31.7) | 172,995 (26.0) | 2,268 (57.0) | 175,263 (26.2) |
| Cardiogenic shock N (%) | 2014 | 13,667 (6.2) | 36,435 (6.0) | 448 (9.3) | 36,883 (6.0) |
| | 2015 | 13,972 (6.4) | 37,725 (6.0) | 470 (9.4) | 38,195 (6.0) |
| | 2016 | 14,175 (6.5) | 39,480 (6.1) | 405 (9.7) | 39,885 (6.1) |
| | 2017 | 14,689 (6.7) | 40,610 (6.1) | 394 (10.0) | 41,004 (6.2) |
| | 2018 | 14,618 (6.9) | 42,280 (6.4) | 380 (10.1) | 42,660 (6.4) |
| | 2019 | 14,137 (6.6) | 43,600 (6.6) | 331 (8.3) | 43,931 (6.6) |
| Resuscitation N (%) | 2014 | 12,118 (5.5) | 11,470 (1.9) | 2,476 (51.7) | 13,946 (2.3) |
| | 2015 | 12,185 (5.6) | 11,735 (1.9) | 2,487 (49.9) | 14,222 (2.2) |
| | 2016 | 12,178 (5.6) | 11,530 (1.8) | 2,141 (51.5) | 13,671 (2.1) |
| | 2017 | 12,112 (5.6) | 11,855 (1.8) | 2,120 (54.0) | 13,975 (2.1) |
| | 2018 | 11,741 (5.5) | 11,925 (1.8) | 1,982 (52.7) | 13,907 (2.1) |
| | 2019 | 11,464 (5.4) | 13,125 (2.0) | 2,106 (52.9) | 15,231 (2.3) |
| Percutaneous coronary intervention N (%) | 2014 | 122,222 (55.7) | 287,470 (47.2) | 287 (6.0) | 287,757 (46.9) |
| | 2015 | 124,489 (56.8) | 296,425 (47.1) | 273 (5.5) | 296,698 (46.7) |
| | 2016 | 128,586 (58.7) | 306,095 (47.0) | 285 (6.9) | 306,380 (46.8) |
| | 2017 | 129,821 (59.6) | 311,030 (47.0) | 310 (7.9) | 311,340 (46.7) |
| | 2018 | 129,419 (60.9) | 313,440 (47.6) | 284 (7.5) | 313,724 (47.4) |
| | 2019 | 131,960 (62.1) | 320,980 (48.3) | 274 (6.9) | 321,254 (48.0) |
| Coronary artery bypass graft N (%) | 2014 | 12,166 (5.5) | 52,820 (8.7) | <=10 (<=0.2) | 52,825 (8.6) |
| | 2015 | 12,042 (5.5) | 54,815 (8.7) | 0 (0.0) | 54,815 (8.6) |
| | 2016 | 12,174 (5.6) | 56,945 (8.7) | 0 (0.0) | 56,945 (8.7) |
| | 2017 | 12,139 (5.6) | 57,420 (8.7) | 0 (0.0) | 57,420 (8.6) |
| | 2018 | 11,608 (5.5) | 57,420 (8.7) | 0 (0.0) | 57,420 (8.7) |
| | 2019 | 11,468 (5.4) | 59,450 (8.9) | 0 (0.0) | 59,450 (8.9) |

### Supplementary material 3 (continued)

| | Year | Germany (full sample) | United States (estimates weighted to national average) | | |
|---|---|---|---|---|---|
| | | DRG statistics | National Inpatient Sample (NIS) | Nationwide Emergency Department Sample (NEDS) | NIS and NEDS combined |
| | | *Inpatient treatment (including ED deaths)* | *Inpatient treatment* | *ED death without admission* | *Inpatient treatment or ED death without admission* |
| Transferred-in from other acute care hospital N (%) | 2014 | 31,902 (14.5) | 107,265 (17.5) | n/a | 107,265 (17.5) |
| | 2015 | 31,319 (14.3) | 109,810 (17.3) | n/a | 109,810 (17.3) |
| | 2016 | 30,144 (13.8) | 113,625 (17.4) | n/a | 113,625 (17.3) |
| | 2017 | 29,397 (13.5) | 118,665 (17.9) | n/a | 118,665 (17.8) |
| | 2018 | 27,241 (12.8) | 120,705 (18.3) | n/a | 120,705 (18.2) |
| | 2019 | 25,695 (12.1) | 125,150 (18.8) | n/a | 125,150 (18.7) |
| Treated in emergency department N (%) | 2014 | n/a | 425,110 (69.8) | 4,790 (100.0) | 429,900 (70.1) |
| | 2015 | n/a | 445,075 (70.7) | 4,981 (100.0) | 450,056 (70.9) |
| | 2016 | n/a | 464,840 (71.4) | 4,162 (100.0) | 469,002 (71.6) |
| | 2017 | n/a | 480,645 (72.6) | 3,924 (100.0) | 484,568 (72.8) |
| | 2018 | n/a | 494,000 (75.0) | 3,763 (100.0) | 497,764 (75.2) |
| | 2019 | n/a | 502,265 (75.5) | 3,978 (100.0) | 506,243 (75.7) |
| Same day stay N (%) [b] | 2014 | 14,097 (6.4) | 18,010 (3.0) | 4,790 (100.0) | 22,800 (3.7) |
| | 2015 | 14,163 (6.5) | 18,370 (2.9) | 4,981 (100.0) | 23,351 (3.7) |
| | 2016 | 14,125 (6.4) | 19,700 (3.0) | 4,162 (100.0) | 23,862 (3.6) |
| | 2017 | 13,919 (6.4) | 20,655 (3.1) | 3,924 (100.0) | 24,579 (3.7) |
| | 2018 | 13,124 (6.2) | 19,860 (3.0) | 3,763 (100.0) | 23,623 (3.6) |
| | 2019 | 12,605 (5.9) | 21,415 (3.2) | 3,978 (100.0) | 25,393 (3.8) |
| Length of stay Median (P25 - P75) | 2014 | 6 (3 - 10) | 2 (1 - 5) | n/a | 2 (1 - 5) |
| | 2015 | 6 (3 - 9) | 2 (1 - 5) | n/a | 2 (1 - 5) |
| | 2016 | 6 (3 - 9) | 2 (1 - 5) | n/a | 2 (1 - 5) |
| | 2017 | 6 (3 - 9) | 2 (1 - 5) | n/a | 2 (1 - 5) |
| | 2018 | 5 (3 - 9) | 2 (1 - 5) | n/a | 2 (1 - 5) |
| | 2019 | 5 (3 - 9) | 2 (1 - 5) | n/a | 2 (1 - 5) |
| Transferred-out to other acute care hospital N (%) | 2014 | 39,145 (17.8) | 49,550 (8.1) | n/a | 49,550 (8.1) |
| | 2015 | 38,229 (17.4) | 50,170 (7.9) | n/a | 50,170 (7.9) |
| | 2016 | 37,036 (16.9) | 50,875 (7.8) | n/a | 50,875 (7.8) |
| | 2017 | 36,125 (16.6) | 50,905 (7.7) | n/a | 50,905 (7.6) |
| | 2018 | 33,517 (15.8) | 48,905 (7.4) | n/a | 48,905 (7.4) |
| | 2019 | 31,676 (14.9) | 48,275 (7.3) | n/a | 48,275 (7.2) |

**Supplementary material 3 (continued)**

| | Year | Germany (full sample) | United States (estimates weighted to national average) | | |
|---|---|---|---|---|---|
| | | DRG statistics | National Inpatient Sample (NIS) | Nationwide Emergency Department Sample (NEDS) | NIS and NEDS combined |
| | | *Inpatient treatment (including ED deaths)* | *Inpatient treatment* | *ED death without admission* | *Inpatient treatment or ED death without admission* |
| In-hospital death N (%) | 2014 | 18,853 (8.6) | 30,315 (5.0) | 4,790 (100.0) | 35,105 (5.7) |
| | 2015 | 18,680 (8.5) | 30,205 (4.8) | 4,981 (100.0) | 35,186 (5.5) |
| | 2016 | 17,858 (8.1) | 30,700 (4.7) | 4,162 (100.0) | 34,862 (5.3) |
| | 2017 | 17,936 (8.2) | 31,035 (4.7) | 3,924 (100.0) | 34,959 (5.2) |
| | 2018 | 17,533 (8.3) | 30,150 (4.6) | 3,763 (100.0) | 33,913 (5.1) |
| | 2019 | 16,881 (7.9) | 29,935 (4.5) | 3,978 (100.0) | 33,913 (5.1) |
| In-hospital death within 30 days N (%) | 2014 | 18,307 (8.3) | 29,905 (4.9) | 4,790 (100.0) | 34695 (5.7) |
| | 2015 | 18,146 (8.3) | 29,760 (4.7) | 4,981 (100.0) | 34741 (5.5) |
| | 2016 | 17,384 (7.9) | 30,265 (4.6) | 4,162 (100.0) | 34,427 (5.3) |
| | 2017 | 17,406 (8.0) | 30,620 (4.6) | 3,924 (100.0) | 34,544 (5.2) |
| | 2018 | 17,106 (8.1) | 29,725 (4.5) | 3,763 (100.0) | 33,488 (5.1) |
| | 2019 | 16,482 (7.8) | 29,455 (4.4) | 3,978 (100.0) | 33,433 (5.0) |

AMI: acute myocardial infarction; ED: emergency department; n/a: not available. [a] Directly standardized by sex and 5-year age-groups according to the 2010 OECD standard population, age >=15. [b] As in US NEDS data no information on length of stay is available for ED visits without inpatient admission, ED deaths without admission were assigned to a length of stay <1 day (same-day stay).
